# Knowledge, Awareness, and Health-Seeking Behaviour Regarding Urinary Schistosomiasis Among Female Adolescents in Riverine Communities of Abuja, Nigeria

**DOI:** 10.64898/2026.01.02.26343347

**Authors:** Opeyemi Adedapo Adetayo, Abubakar Imam, Akolo Yohanna Jaggu, Unekwuojo Iye Etubi-Ibrahim, Olubunmi Iyabode Ojji, Bibiana Nonye Egenti

## Abstract

**Background:** The success of programs to control Neglected Tropical Diseases (NTDs) like schistosomiasis is heavily dependent on the knowledge and health behaviors of affected populations. This study aimed to assess the knowledge, awareness, and health-seeking behaviours regarding urinary schistosomiasis among female adolescents, a key demographic, in riverine communities of Abuja, Nigeria.

**Methods:** A descriptive cross-sectional study was conducted among 300 female adolescents (aged 10–19) in Gui Ward, Abuja, selected via a multistage sampling technique. A pre-tested, semi-structured questionnaire was used to assess sociodemographic data and specific knowledge about urinary schistosomiasis, including awareness of the disease, its transmission, symptoms, and prevention. Data were analyzed using descriptive and inferential statistics.

**Results:** The study revealed a profound lack of knowledge. Only 27.3% (82/300) of respondents had ever heard of schistosomiasis. Among those aware, only 14.6% (12/82) knew the correct mode of transmission (contact with contaminated water). A significant misconception was the belief that the disease is transmitted by drinking contaminated water, which was the most frequently mentioned mode of transmission (39%). There was a significant association between this misconception and infection status (p=0.048). Despite low awareness, symptoms were common, but health-seeking from formal healthcare facilities was suboptimal.

**Conclusion:** Limited knowledge and misconceptions about urinary schistosomiasis hinder control efforts among female adolescents in Nigerian communities, increasing vulnerability and transmission. There is an urgent need for targeted health education programs in schools and communities that address local misconceptions and integrate with WaSH improvements and mass drug administration.

## Introduction

Urinary schistosomiasis, caused by Schistosoma haematobium, is a debilitating parasitic disease that disproportionately affects impoverished communities in sub-Saharan Africa [1, 2]. While control strategies, primarily preventive chemotherapy with praziquantel, have been scaled up, their success is often undermined by high rates of reinfection. Sustainable control and eventual elimination of the disease cannot be achieved through mass drug administration (MDA) alone. It requires a comprehensive approach that includes improvements in water, sanitation, and hygiene (WaSH), snail control, and, crucially, behavioral change within at-risk populations [3,4].

There is a growing body of evidence demonstrating a direct link between low knowledge, negative attitudes, and risky WaSH practices and the continued transmission of schistosomiasis [5,6]. The perception of schistosomiasis as a minor illness, rather than a serious threat to health and development, contributes to the persistence of the disease. Therefore, understanding a community’s knowledge, attitudes, and practices (KAP) is a critical prerequisite for designing and implementing effective behavioral change interventions that promote the adoption of protective behaviors and the reduction of risk [7].

Adolescents, particularly girls, are a key demographic in the transmission cycle. They often bear the responsibility for domestic chores like washing clothes and fetching water, which puts them in frequent contact with potentially contaminated water sources [8]. Furthermore, they are at an age where the long-term consequences of infection, such as female genital schistosomiasis (FGS), can have a devastating impact on their reproductive health and social well-being [9,10]. Despite their vulnerability, their knowledge and perceptions of the disease are often poorly understood, as control programs have historically focused on younger, school-aged children.

Studies across sub-Saharan Africa have consistently revealed significant gaps in knowledge. A survey in Mozambique found that while general awareness of schistosomiasis was high (91%), correct knowledge of its transmission and prevention was very low (18% and 13%, respectively) [11]. Similarly, a qualitative study in Ghana found that knowledge of FGS was almost non-existent among women, girls, and even frontline health workers, with symptoms often being misattributed to sexually transmitted infections (STIs), leading to stigma and inappropriate treatment [8,12]. This misperception is particularly damaging, as it can deter individuals from seeking timely and appropriate care.

This study was designed to address this gap by specifically assessing the knowledge, awareness, and health-seeking behaviors related to urinary schistosomiasis among female adolescents in riverine communities of Abuja, Nigeria. The aim was to identify the specific knowledge gaps and misconceptions that fuel vulnerability in this group, in order to provide evidence for the development of targeted and effective health education interventions.

## Methods and Materials

### Study Setting

The study was conducted in Kuyami Resettlement village and Gosa-Kpayin-kpayin, two riverine communities in Gui Ward of the Abuja Municipal Area Council (AMAC), Nigeria. These communities were selected due to their reliance on natural water bodies for daily activities.

### Study Population

The study population comprised female adolescents aged 10 to 19 years.

### Study Design and Sample size

A descriptive cross-sectional study design was used. The sample size was calculated Leslie Kish formula for sample size estimation giving a sample size of 300 participants.

### Sampling Procedure

A multistage sampling technique was used to select the Area Council, Ward, and riverine communities, clusters and adolescent girls.

### Data Collection Instrument

A pre-tested, semi-structured questionnaire, adapted from a previous study [2], was used for data collection. The questionnaire was administered by trained research assistants in the participants’ preferred language (English, Hausa, Gbagyi, or Pidgin English). The key section for this study was Section C: History of Urinary Schistosomiasis, which was designed to assess:

#### Awareness

Whether the participant had ever heard of the disease.

#### Knowledge of Symptoms

Participants were asked to name symptoms they associated with the disease.

#### Knowledge of Transmission

Participants who were aware of the disease were asked how a person could get it. Responses were recorded and later categorized.

#### Knowledge of Prevention

Participants were asked how the disease could be prevented.

#### Health-Seeking Behaviour

Participants were asked what action they would take or had taken if they experienced symptoms like blood in urine.

### Data Analysis

Data was analyzed using IBM SPSS statistics software Version 29. Descriptive statistics (frequencies and percentages) were used to summarize the levels of awareness and knowledge. The Chi-squared test was used to assess the association between knowledge (and misconceptions) and infection status, which was determined through urine microscopy as described in the companion paper. A p-value < 0.05 was considered statistically significant.

### Ethical Considerations

Ethical approval was obtained from the Health Research Ethics Committee of the Federal Capital Development Authority (FHREC/2022/01/221/07-11-22). Community entry protocols were followed, and informed consent/assent was obtained from all participants and their guardians. Confidentiality was maintained, and all infected individuals received free treatment free of charge.

## Results

**Table 1:** Sociodemographic Characteristics of Respondents.

| Variable | Frequency (n=300) | Percentage (%) |
| --- | --- | --- |
| **Community** |  |  |
| Kuyami | 157 | 52.3 |
| Gosa-Kpayinkpayin | 143 | 47.7 |
| **Age Group (years)** |  |  |
| 10-14 | 196 | 65.3 |
| 15-19 | 104 | 34.7 |
| **Religion** |  |  |
| Christianity | 227 | 75.7 |
| Islam | 73 | 24.3 |
| **Education Level** |  |  |
| Primary | 141 | 47.0 |
| Secondary | 150 | 50.0 |
| Tertiary | 3 | 1.0 |
| Out of school | 6 | 2.0 |

### Awareness of Urinary Schistosomiasis

The overall awareness of urinary schistosomiasis was exceptionally low. Majority of the respondents, 218 out of 300 (72.7%), indicated that they had never heard of a disease called schistosomiasis, even when local names were used. Only 82 respondents (27.3%) were aware of the disease.

Crucially, the misconception that schistosomiasis is transmitted through drinking dirty water was found to be significantly associated with having the infection (p=0.048). This suggests that this specific misunderstanding may lead individuals to believe they are safe as long as they do not drink stream water, while they continue to engage in high-risk contact activities.

**Table 2:** Knowledge of Transmission Among Respondents Aware of Schistosomiasis (n=82)

| Mode of Transmission Mentioned | Frequency | Percentage (%) |
| --- | --- | --- |
| Contact with contaminated water (Correct) | 12 | 14.6 |
| Drinking contaminated water (Incorrect) | 32 | 39.0 |
| Wading in dirty water (Correct) | 5 | 6.1 |
| Don't know | 27 | 32.9 |
| Other (e.g., witchcraft, flies) | 6 | 7.3 |

### Knowledge of Symptoms and Health-Seeking Behaviour

In contrast to the poor knowledge of transmission, awareness of symptoms was comparatively higher. Among those who had heard of the disease, 58 (70.7%) could name at least one symptom. The most commonly mentioned symptom was genital itching (27%), followed by blood in urine (haematuria) (26.7) and painful urination (17%).

However, knowledge of symptoms did not translate to appropriate health-seeking behaviour. This study noted that among all participants who reported having experienced haematuria in the past (81), only 15(18.8%) had sought treatment from a formal health facility and only 10 (12.3%) actually received treatment. Others reported seeking no treatment, using traditional remedies, or visiting a patent medicine vendor.

## Discussion

The findings of this study reveal a critical and dangerous knowledge gap regarding urinary schistosomiasis among adolescent girls in the study communities. The fact that nearly three-quarters of the respondents had never heard of the disease, which is endemic in their area, is a major failure of public health communication and a significant barrier to control. This finding is more alarming than those from studies in Mozambique and Ghana, where general awareness was high, even if specific knowledge was lacking [11,13]. It indicates that in these Nigerian communities, schistosomiasis is truly a “neglected” disease, not just for policymakers, but for the very people who suffer from it.

The most damaging finding is the widespread and statistically significant misconception that the disease is transmitted through drinking contaminated water. This fundamental misunderstanding of the transmission pathway means that even if individuals are aware of the disease, their preventive actions are likely to be ineffective. They may focus on securing safe drinking water while failing to avoid skin contact with the same water sources during bathing, laundry, or agricultural work; the very activities that pose the highest risk. This directly explains why, as found in the companion epidemiological study, water contact is a primary driver of the 39.7% prevalence in this same population.

This study’s results are consistent with a systematic review of KAP studies in sub-Saharan Africa, which found that limited knowledge and risky water practices are key factors promoting transmission [6]. The reluctance to seek formal medical care, with only half of those with haematuria visiting a health facility, is also a major concern. This may be partly due to the stigma identified in other studies, where symptoms like genital itching and discharge are misperceived as STIs [8,12]. This can lead to shame, social isolation, and a reluctance to seek care, allowing the disease to progress to more severe forms like FGS, with devastating long-term consequences for reproductive health.

The clear implication is that MDA campaigns, while essential, are insufficient on their own. Without addressing the fundamental lack of knowledge and correcting dangerous misconceptions, reinfection rates will remain high, and the cycle of transmission will continue. There is an urgent need for school- and community-based health education. Schools are a powerful channel for reaching adolescents, who can then act as “change agents” within their families and communities [7]. Health education must be tailored to the local context, using local languages and directly addressing the specific misconceptions identified in this study.

## Data Availability

All data produced in the present study are available upon reasonable request to the authors

## Declarations

### Ethical Approval and Accordance

Ethical approval for this study was obtained from the Health Research Ethics Committee of the Federal Capital Development Authority. (Approval Number: **FHREC/2022/01/221/07-11-22**). The study was conducted in full accordance with the guidelines, regulations, and ethical standards set forth by the ethics committee.

### Consent to Participate

Written informed consent was obtained from all participants before enrolment into the study. Participation was voluntary, and confidentiality was assured.

### Consent to Publish

Not applicable: No identifying information or images of individual participants are included in this manuscript.

## Acknowledgement

The authors acknowledge the Network on Behavioral Research for Child Survival (NETBRECSIN) for reviewing this work and offering valuable feedback that enhanced this study

## Data Availability Statement

The datasets generated and analysed during this study contain sensitive personal information and are therefore not publicly available in order to protect participant confidentiality. However, the identified data may be made available from the corresponding author upon reasonable request and subject to approval by the relevant ethics committee.

## Conclusion

Low knowledge of urinary schistosomiasis is a primary driver of vulnerability among female adolescents in these Nigerian riverine communities. The profound lack of awareness and the prevalence of critical misconceptions about the disease’s transmission pathway directly contribute to high-risk behaviors and sustained infection. This study demonstrates that without targeted and sustained health education, the impact of other control measures will be severely limited. It is imperative that public health authorities and their partners develop and implement community-based educational campaigns that are integrated with WaSH improvements and MDA programs. Empowering adolescent girls with the correct knowledge to protect themselves is not just a component of schistosomiasis control; it is the foundation upon which all other efforts must be built.

